# Mechanistic modeling-guided optimization of microneedle-based skin patch for rapid transdermal delivery of naloxone for opioid overdose treatment

**DOI:** 10.1101/2022.03.18.22272612

**Authors:** Akeemat Tijani, Prashant Dogra, Maria J. Peláez, Zhihui Wang, Vittorio Cristini, Ashana Puri

## Abstract

Naloxone, an FDA approved opioid inhibitor, used to reverse opioid overdose complications has up till date faced challenges associated with its delivery. Limitations include the use of invasive delivery forms and the need for frequent redosing due to its short half-life. Further, the use of the non-invasive commercially available intranasal form is faced with limitations such as nasal epistaxis and inability to use the intranasal channel due to nasal trauma common in addicts from frequent drug snorting. The goal of the current study was to design a rapidly dissolving polymeric microneedle (MN) patch with delivery and pharmacokinetic properties comparable to that seen with the commercially available NAL products, eliminating their highlighted limitations. Factors such as drug loading and polymer strength influenced the fabrication of the MNs. Compared to passive permeation, a reduced lag time of about 5-15 min was observed with a significant drug flux of 15.09 ± 7.68 μg/cm^2^/h seen in the first 1 h (p<0.05) with an array of 100 needles (500 µm long). Increasing the MN length and density (no. of needles/unit area) made a significant difference in the amount permeated and flux (p<0.05), with latter being more influential. Further, through a mechanistic model of drug release from the patch, integrated with pharmacokinetic modeling, we optimized the design of the patch to reproduce in-silico the clinical pharmacokinetics of NAL obtained through commercial intramuscular and intranasal devices. Model simulations and analyses revealed the significance of needle base diameter and needle count in improving systemic pharmacokinetics of NAL.

## 1. Introduction

Opioid use disorder is an agelong societal menace affecting over 16 million people in the world [1]. This includes both prescription and non-prescription misuse of synthetic and natural opioids such as fentanyl, heroin, morphine etc. Death due to overdosage is quite prevalent with opioid abuse [2–5]. It is important to note that at the global level, over a 100,000 opioid overdose related deaths are reported annually with 56,064 deaths reported in the USA alone in 2020 by the CDC’s National Center for Health Statistics [6,7]. This detrimental effect of excessive consumption of opioids has been shown to be mainly due to the agonistic effect of opioids on the mu receptor, which results in significant respiratory depression and various other complications that eventually warrants urgent medical intervention [5,8].

Interestingly, the opioid menace is a global epidemic that has consistently triggered research into the discovery of medications for curbing and managing the disorder. A breakthrough therapeutic for the management of opioid overdosage emergencies is naloxone (NAL), approved by the FDA in 1971 and commonly sold under the brand name, Narcan [9,10]. Challenges encountered with the successful use of this drug include drug delivery limitations. It is currently administered via the intravenous (IV), intramuscular (IM), subcutaneous (SC), and intranasal (IN) route. With IV delivery, even though a desired rapid response is seen within 2 min of drug administration [11], the rapid systemic clearance of the drug and hence its short half-life of about 30-81 min oftentimes warrants repeat dosing which is usually recommended to be given every 2-3 min depending on individual recovery seen and severity of overdosage [10,12,13]. Typically, [10] from a patient perspective, this is deemed not an ultimately preferred option considering the invasive nature of IV delivery, causing needless pain and discomfort through the course of patient stabilization. Further, safety issues relating to inflammation and damage to blood vessels from prolonged drug delivery via the IV route is prevalent [14,15]. It is also a common challenge to have difficulty accessing the vasculature for IV injections since most abusers have their veins damaged from chronic self-drug administration [11,13]. This is currently regarded an avoidable situation considering various successes that have been recorded with respect to drug delivery approaches and drug carriers that have been unveiled by years of research in drug delivery. The need for trained medical intervention at all times seems not so appropriate as well for a drug set for use in emergencies, preferably, this should be available in a form ready for use by lay individuals or first responders in overdose situations [4,11]. Further, care givers are at risk of deadly communicable infections such as HIV/AIDS or hepatitis via accidental needle stick injuries [16].

Oral delivery of NAL on the other hand is impractical due to its significant hepatic metabolism and consequently poor oral bioavailability of about 2% [17]. Efforts to develop alternative non-invasive delivery options brought in its wake the approval of NAL intranasal (IN) spray [4,11]. This is however not devoid of limitations as well and some of these include the inability to use route due to injuries to the nasal mucosa common in drug addicts from drug snorting [13,18]. Response variation among users due to nasal congestion, epistaxis and infection is also a challenge [11,13]. Efforts to develop transdermal formulations for NAL has been previously reported, justification for this being its non-invasiveness and capability for sustained release and minimized side effects. One of such attempts was the development of a reservoir transdermal patch by Panchagnula and his group [19] and also the application of iontophoresis to facilitate skin delivery of NAL by Yamamoto and his coworkers [11,20]. These approaches though quite promising as well considering the attributes of the delivery route are not considered optimal options because what is mostly desired in NAL overdose emergency situations is rapid drug delivery to reverse significant respiratory depression which is the leading cause of death in opioid overdose situations [11].

In the light of this, a fast-rising approach, that is gaining public embrace for the delivery of biopharmaceuticals which is considered a feasible option for solving the problem at hand is the use of microneedles (MNs) for systemic delivery through the skin [21]. This offers numerous advantages that eliminates or covers for most of the highlighted limitations of the current delivery forms of NAL [21–23]. It is predicted considering its ability to create microchannels in skin to be able to offer rapid delivery quite close to that seen with the current delivery routes and systems for NAL and at the same time provide some sustained release effect that will limit the frequency of injections [24–26]. Further, it is anticipated to be considered more favorable to patients since it is minimally invasive, and typically eliminates the need for intervention by a specialized clinician [22,27].

Preliminary studies from our lab have previously demonstrated the effectiveness of the use of solid MNs for the delivery of NAL across the skin, where a short lag time of about 8 min was observed with significant increase in drug permeation seen when compared to passive delivery in an in vitro set up [13]. The feasibility of achieving a quite similar pharmacokinetic (PK) profile to the FDA approved IM and IN formulations by the use of MNs was also predicted by our group through in vitro-in vivo PK modeling [13]. However, from a clinical standpoint, a dissolvable or biodegradable MN patch would be preferred as it would eliminate the poke and patch step typical of solid MNs [26]. For this approach, drug is entrapped in a dissolving or biodegradable polymer, and is released upon insertion in skin, and further diffuses into skin with polymer dissolution [28–30]. With this, there is no need for the needle withdrawal step prior to drug application or even with drug application as is common with coated solid MNs [31]. Dissolvable polymers are adaptable to easy and inexpensive fabrication processes [32]. They are expected to shorten drug delivery time and hence onset of action. Moreover, safety issues including needle breakage in skin and the need for the disposal of needles post usage into the environment which are typical of solid type MNs are evaded [33–35]. Their typical less complex fabrication requirements are expected to be more adaptable to the large-scale use of NAL and the clinical and commercial success of rapidly dissolving NAL MNs are thus foreseen. It is interesting that quite a number of successes have been seen with polymeric MNs for rapid delivery of biopharmaceuticals such as insulin and contraceptives in the literature [25,28,36].

The design of a successful polymeric MN patch involves the careful selection of a starting polymer compatible with the drug under consideration which would produce a final product with strong mechanical properties and at the same time excellent drug permeation efficiency. Based on the requirement for NAL, the rapidly dissolving polymer, polyvinylpyrrolidone (PVP) with significant successes recorded for the encapsulation and microfabrication of MNs for a number of biopharmaceuticals was considered for fabricating NAL MNs. Beyond its rapid dissolving property, its excellent biophysical properties ranging from its biocompatibility to its adaptability to simple microfabrication processes makes it an excellent selection for NAL. It could be used alone without the inclusion of organic solvents [32]. Important considerations for the development of a viable final product include drug loading capacity of the polymer and polymer strength [28]. In the present study, PVP-based NAL MNs were fabricated. Optimal fabrication processes required for the formulation of consistently efficient NAL MNs in terms of penetration and permeation efficiency through skin was determined. To our knowledge, this is so far the first study to fabricate MN skin patches for NAL transdermal delivery. Herein, the optimized microfabrication process for NAL PVP MN, its permeation profile, MN optimization considerations and characterization reports on the fabricated MNs are reported.

Of note, we developed a mechanistic mathematical model to simulate the dissolution of conical MNs and study drug release from dissolvable MN-based patches. For our work, we adapted previously published mathematical models that described the dissolution process of MNs to predict the delivery of drugs into plasma. Kim et.al. developed a model for a conical microneedle from which a surrounding shell dissolved at every timepoint to quantify the dissolved microneedle volume kinetics. Similarly, Zoudani and Soltani et al. implemented a mathematical model to understand the dissolution process of conical microneedles in porous medium, using the same shell dissolving process as above, and incorporated an array of hemispherical convexities loaded with drug on the tip of the microneedle to decrease the drug delivery time [37,38]. In the current study, we assumed a modified dissolution process such that instead of the needles shrinking in thickness due to dissolution, we assumed shrinkage of length as they dissolve, to better reflect the observations in the literature [39,40]. Further, based on our previous works [13], by integrating to a two-compartment PK model, our model can predict the clinical PK of NAL. Through comprehensive parameter analyses, the model was thus used to optimize the design of patches to improve drug release characteristics and enhance plasma bioavailability of NAL to match the clinical performance of FDA-approved devices for NAL delivery.

## 2. Materials

NAL hydrochloride was purchased from Thermo Fisher Scientific (Ward Hill, MA, USA). Polydimethylsiloxane (PDMS) molds were obtained from MicroPoint Technologies (Pioneer Junction, Singapore). Trifluoroacetic acid (TFA), potassium monophosphate, 10X phosphate buffered saline, pH 7.4 (PBS), PVP and silver wire (0.5 mm diameter, 99.99%) were purchased from Fisher Scientific (Fair lawn, NJ, USA). Methanol was purchased from Concord Technology (Beichen, Tianjin, China). Silver/silver chloride electrodes (2 mm ×4 mm) were obtained from A-M systems (Sequim, WA, USA). Flourescein isothiocyanate was bought from Sigma Aldrich (St. Louis, MO, USA). Methylene blue was obtained from Electron Microscopy Sciences (Hatfield, PA, USA). Porcine ears were procured from Animal Technologies (Tyler, Texas, USA).

## 3. Methods

### 3.1. Solubility study

The solubility of NAL in 7.5% PVP and 10 mM PBS, pH 7.4 (1X PBS) was determined. For all, an excess amount of NAL was added to 200 µL of these solvents and left for shaking (Orbi-Shaker BT302, Cambridge Scientific, Boston, MA) at room temperature for 24 h. The solution was then centrifuged (05-090-128 Mini Centrifuge, Fisher Scientific, Fair Lawn, NJ) and the supernatant was diluted 10,000 times with 1X PBS and analyzed using reverse phase high performance liquid chromatography (RP-HPLC, n = 3) [13].

### 3.2. Fabrication of NAL MNs

NAL loaded MNs with varying dimensions were prepared with details shown in **Table 1**. This was done using the mold casting technique with PDMS molds. A known amount of drug was dissolved in 7.5% solution of PVP in distilled water to prepare casting solutions of varying drug strength ranging from 50 through 150 mg/ml of NAL in the preformed PVP solution. The drug solution (170 µL) was poured in the molds and vacuumed at room temperature and a constant pressure of about -0.6 psi for 5 min with iterative degassing steps. Following the full elimination of air bubbles in mold filled solutions, 130 µL of drug in 7.5% PVP solution was added. Full degassing was further ensured and vacuum with same condition as starting steps was applied for at least 1 h and left for drying. The mold was filled up to its brim after 24 h with 130 µL of drug in 7.5% PVP solution to strengthen and thicken the base of the needle. The needles were left in the desiccator for drying at room temperature for two more days. Following this, needles were gently separated from the mold and transferred onto a 3M adhesive tape backing for application on skin. The mold casting technique is illustrated in **Fig. 1**.

**Table 1.**
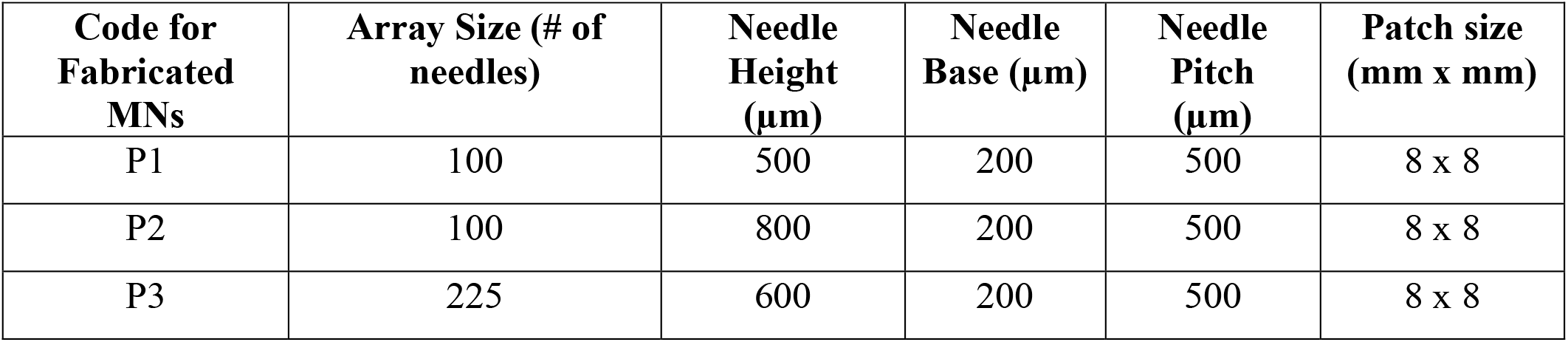
Dimensions of fabricated NAL MN patches.

| Code for Fabricated MNs | Array Size (# of needles) | Needle Height (μm) | Needle Base (μm) | Needle Pitch (μm) | Patch size (mm x mm) |
| --- | --- | --- | --- | --- | --- |
| P1 | 100 | 500 | 200 | 500 | 8 x 8 |
| P2 | 100 | 800 | 200 | 500 | 8 x 8 |
| P3 | 225 | 600 | 200 | 500 | 8 x 8 |

**Fig. 1.**
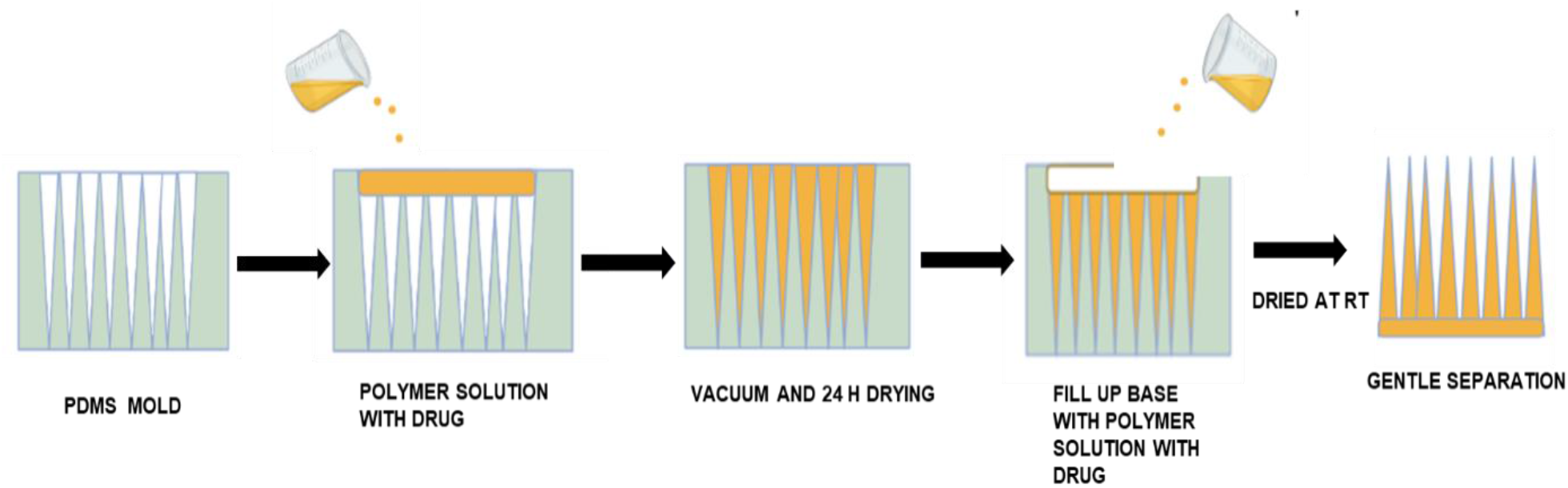
Schematic illustration of the mold casting procedure followed for fabrication of NAL PVP-based MNs

### 3.3. Characterization of NAL MNs

#### 3.3.1. Bright field imaging

Imaging was done using the Omax brightfield microscope (OMAX MD82EZ10, China). A sample holder was manually formed in the lab to fix MNs on the microscope stub. This was done using double sided tapes, microscope glass slides and a parafilm bed to hold the MNs at an angle suitable for viewing it sideways. This was used to evaluate MNs for crystals, qualitative and quantitative assessments including the evaluation of needles for surface morphology, needle tip integrity, uniformity and dissolution post-insertion for 2 min. All these were determined by running across the entire needle array using the 40X magnification lens. Images were taken using the Toup view imaging software. Calibration was done with a 1 mm rule to measure MN length, tip to tip distance, tip to base and base to base distance (n = 6). Results are presented as mean ± SE and compared to the vendors specifications.

#### 3.3.2. Dye binding and skin insertion efficiency

Skin pieces were obtained and thawed, dabbed with Kim wipes to remove moisture from thawing. MNs were applied using thumb insertion force for 2 min. Thereafter, a drop of methylene blue dye was applied for about 20 min. Dye on skin was wiped off with Kim wipes and skin was gently cleaned with alcohol swabs. Evaluation of the stained skin for pore formation was done under the microscope [12]. Further, skin insertion efficiency was determined by counting the number of pores per porated skin (n=3-4) and reporting as a percentage based on the total number of needles on the array mold provided by the vendor ± SE.

#### 3.3.3. Confocal laser scanning microscopy

The depth of the microchannels created by MNs was analyzed using confocal laser scanning microscopy (CLSM). Fluorescein isothiocyanate (0.35%, 200 µL) was applied to the skin after treatment with MNs for 1 min. Excess dye was removed with Kimwipes. The MN treated skin samples were placed on a glass slide without distortion or fixation artifacts. The slides were observed using a computerized Leica TCS SPE II confocal microscope (Leica microsystems, Heerbrugg, CH9435 Switzerland) with 10X objective at an excitation wavelength of 488 nm. Leica Application Suite X 3.5.5.19976 fluorescence software was used to process the images. The depth of the microchannels created and pattern of distribution of fluorescein in the microchannels was studied using X-Z sectioning [13].

#### 3.3.4. Drug content

Fabricated MN arrays (n = 4) were separated gently, dissolved in 20 ml of 1X PBS and vortexed. The solutions were then diluted 100 times with 1X PBS and HPLC was used to quantitate the drug content in each diluted solution. The quantity of drug in each sample ± SE was determined by the standard curve method.

### 3.4. *In vitro* permeation study

Porcine ear skin was trimmed and dermatomed, cut into appropriate pieces (thickness ranging between 0.6-0.9 mm) and stored in a freezer at -20^0^ C. Skin pieces were removed on the day of the study and thawed in 1X PBS solution (pH 7.4) at 37°C for 2-3 min. Receptor for Franz cell diffusion chambers were filled up with 5 ml of 1X PBS. Prepared skin pieces were mounted in between the receptor and donor. The donor chambers were filled with 300 µL of 1X PBS. After about 15 min of equilibration, skin integrity was determined by measuring skin resistance. Silver/silver chloride electrodes, an arbitrary 2 channel waveform generator (RIGOL DG822, 25 MHZ, TEquipment, NJ, USA) and a digital micrometer (RIGOL DM 3068 61/2 digit, TEquipment, Long Branch, NJ, USA) were used for this. The load resistor R_L_ (silver chloride, 100 kΩ) was dipped in the donor solution, The value on the multimeter was recorded. Skin pieces with display values ranging from 15-50 mV were selected for the studies. Resistance was then calculated using the formula:

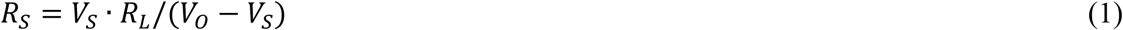

Where V_S_ is the voltage drop across the entire circuit, R_L_ is load resistance and V_O_ is voltage drop across skin. V_O_ and R_L_ were 100 mV and 100 kΩ, respectively. Care was taken to ensure that the resistances of the skin pieces within the groups all through studies were comparable. After measuring the resistance, the donor solution was removed, and dried with Kim wipes. Skin temperature was measured using IR thermometer ensuring that they were all about 32° C. The MN patches of varying dimensions as detailed in **Table 1** were adhered onto skin using the adhesive backing and thumb force for 2 min was applied for microporation on the selected skin pieces. The skin pieces with the patches adhering to them were then mounted on the Franz cells and left for 24 h. The control group assessed the passive permeation of NAL across intact dermatomed porcine ear skin after applying 430 µL of 50 mg/ml NAL in 7.5% PVP solution in the donor chamber (n = 5). Receptor samples (300 µL) were taken at 0 h, 0.083 h, 0.25 h, 0.5 h, 0.75 h, 1 h, 2 h, 4 h, 5 h, 6 h, 8 h, 22 h, 24 h, into sample vials and filtered. Equal volume of fresh buffer solution was replaced in receptor chamber of Franz cells. Samples were then analyzed for drug amount using HPLC. The result for each test group was reported as mean ± SE. The cumulative amount of NAL permeated through skin per unit area was plotted as a function of time. The slope of the linear portion of the plots over the first hour and over 24 h was used to calculate the flux of NAL from the patch [13,41].

### 3.5. Quantitative analysis of NAL

HPLC analysis for NAL was done using the HPLC Waters Alliance 2695 separation module (Cambridge Scientific, MA, USA). The validated method used was an isocratic elution that involved the use of Kinetex® biphenyl 100 A∘ 250 × 4.6 mm, 5 µm C18 column (Phenomenex, CA, USA). The mobile phase and volume ratios include 30:60:10 v/v methanol with 0.1% TFA, water, and 0.2 M phosphate buffer pH 4.0, respectively set to flow at the rate of 1.05 ml/min. Sample run time was 8 min with retention seen at about 4.7 min. Drug peaks eluted at wavelength 211 nm. A calibration curve was obtained over a drug concentration range of 0.1-50 µg/ml with 1X PBS used as diluent. Linearity was obtained with fitted plot (R^2^ = 0.9999). The limits of quantification (LOQ) and detection (LOD) were determined as 0.13 and 0.04 µg/ml, respectively [13].

### 3.6. Mathematical modeling

#### 3.6.1. Mathematical model development

We adapted the Nernst-Brunner equation [42] (Eq. 2) to model the dissolution kinetics of MN patches and quantify the release of NAL into the Franz cell.

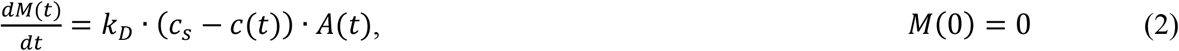

Here, *M*(*t*) is the dissolved mass of the patch; *k*_*D*_ is the dissolution rate constant of the matrix polymer, which is equal to the ratio of diffusion coefficient (*D*) of the polymer to the thickness (*ε*) of the unstirred layer (i.e.,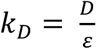); *A*(*t*) is the available surface area of the patch for dissolution; *c*_*s*_ is the solubility of the polymer, and *c*(*t*) is the concentration of the dissolved polymer in bulk solution.

As a simplification, we applied the above equation to a single repeating unit of the patch, defined by a single microneedle and the supporting piece of baseplate (or backing) and thus obtained the cumulative mass kinetics of the entire patch by multiplying over *N* repeating units (**Fig. 2a**). Note that as a further simplification, we assumed *c*(*t*) = 0, suggesting that polymer concentration in the skin is effectively zero due to its rapid transport into the Franz cell solution *in vitro*, or analogously into the systemic circulation in an *in vivo* setting. Therefore, we obtained:

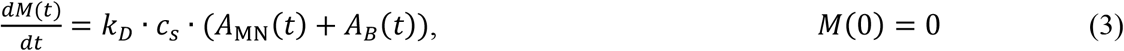

where, *A*_MN_(*t*) and *A*_*B*_(*t*) are the available surface areas of a single microneedle and its associated baseplate, respectively.

**Fig. 2.**
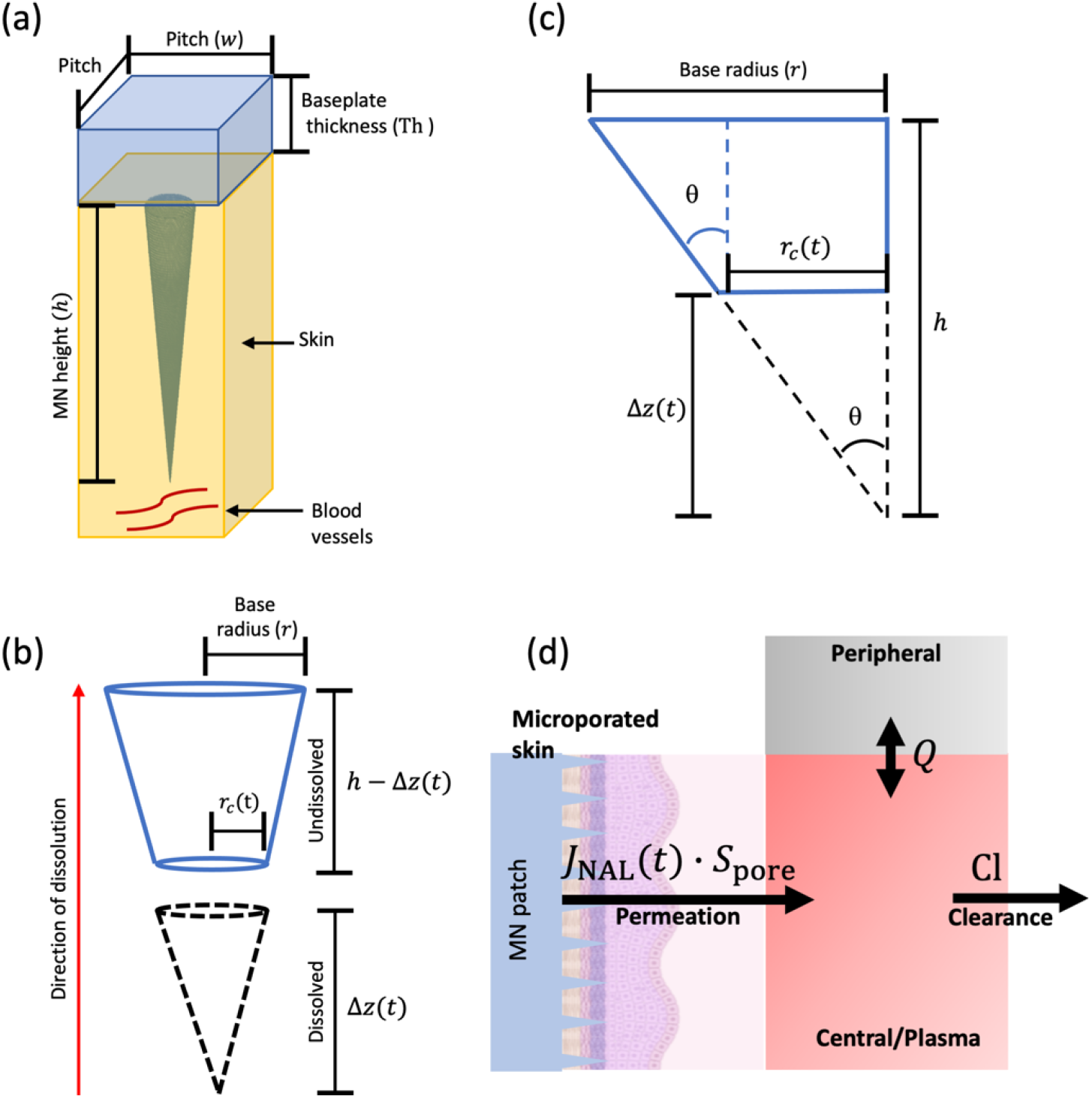
Mathematical modeling. **(a)** Representative repeating unit of the MN patch containing a single MN and the associated baseplate used for model development. (**b)** Schematic of a single MN representing the dissolution process starting at the tip of the cone and moving towards the base. (**c)** Schematic showing trigonometric relationships in the cone. *θ* is the half angle at the apex of the cone. (**d)** Two-compartment pharmacokinetic model integrated to the patch-based drug delivery model.

To estimate the available MN surface area (*A*_MN_(*t*)), we assumed that the MN is shaped like a cone that dissolves starting from its tip and ending at its base **Fig. 2b**, as observed in the literature [43,44]. Therefore, at any timepoint *t*, the MN can be viewed as a truncated cone with surface area available for dissolution given by:

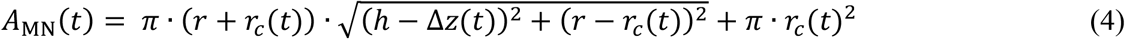

where, *r* and *r*_*c*_(*t*) are the large and small radius of the truncated cone, respectively; *h* is the initial height of the intact MN; and Δ*z*(*t*) is the dissolved height of the MN.

The half angle at the apex of the cone (*θ*) is related to the radius and the height of the intact cone as 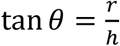, and by similarity of triangles, we can also relate it to the parameters of the truncated cone, such that 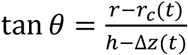 (**Fig. 2c)**. With this relation, we can rewrite Eq. 4 in terms of the MN parameter *θ* as:

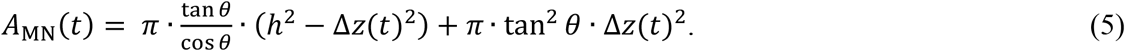

Further, since in the Nernst-Brunner equation we are modeling the dissolved mass *M*(*t*) of the MN, therefore, to retain only one variable in the model, in Eq. 5 we rewrite the dissolved height of the cone Δ*z*(*t*) in terms of the corresponding dissolved mass *M*(*t*). Given the volume of a cone being 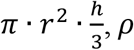, *ρ* as the density of the MN, and 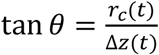 from similarity of triangles (**Fig. 2c)**, we obtain the mass of the dissolved cone:

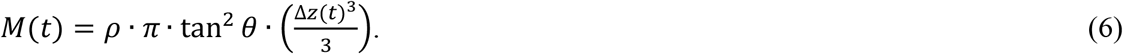

Solving for Δ*z*(*t*) in the above expression and substituting its value in Eq. 5, we obtain:

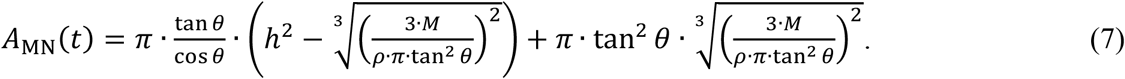

Further, the surface area of dissolution for the associated baseplate of the MN is 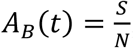, where *S* is the total surface area of the patch and *N* is the number of MNs (or repeating units) in the patch. Substituting this expression and Eq. 7 in Eq. 3, we obtain:

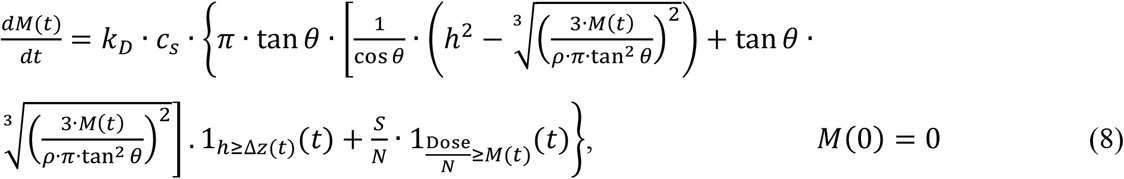

where, 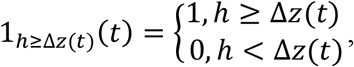,

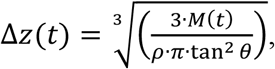

and 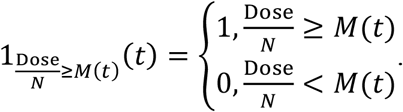

Note that we did not assume any time delay in the initiation of dissolution of baseplate, i.e. the MNs and baseplate begin to dissolve simultaneously upon administration.

By solving the above equation and multiplying with the *N* repeating units of the patch, we obtained the total cumulative mass (*M*(*t*)) of the dissolved patch. From this, we estimated the total cumulative mass of NAL (*M*_NAL_(*t*)) released by the patch into the Franz cell by multiplying with the drug loading fraction *β* of the patch:

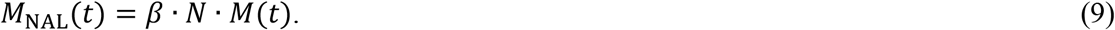

Eq. 9 was numerically solved in MATLAB using the built-in ODE solver *ode45*. Non-linear least squares regression of the model solution to *in vitro* release kinetics data for NAL was performed to estimate the unknown model parameter *k*_*D*_, using the built-in MATLAB function *lsqcurvefit*. The remaining model parameters were known a priori (**Table 2)**. Further, from the numerical solution of the model, the time-dependent flux of NAL (*J*_NAL_(*t*)) was estimated as 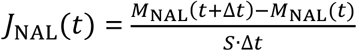, to which spline interpolation was performed to obtain a time-dependent function of flux for application in pharmacokinetic analysis.

**Table 2.** List of model parameters.

| Parameter | Definition | Value |  |  | Units | Ref. |
| --- | --- | --- | --- | --- | --- | --- |
| *Patch-related parameters* |  |  |  |  |  |  |
|  |  | P1 | P2 | P3 |  |  |
| $r^*$ | Base radius of MN | 0.01 | | | cm | Exp. |
| $h^*$ | Height of MN | 0.047 | 0.068 | 0.0561 | cm | Exp. |
| $\theta$ | Half angle of cone apex | 12.01 | 8.37 | 10.1 | Degree | Calc. |
| $N^*$ | Number of MNs in a patch | 100 | 100 | 225 | - | Exp. |
| $\rho^*$ | Material density of patch | 1.5e+6 | | | $\mu\text{g}/\text{cm}^3$ | [46] |
| $c_s^*$ | Solubility of patch matrix | 1.6e+5 | | | $\mu\text{g}/\text{mL}$ | Exp. |
| $w^*$ | Pitch of the patch | 0.0482 | 0.0488 | 0.0476 | cm | Exp. |
| Dose | Dose of drug loaded in the patch | 17895.94 | 17203.9 | 17814.45 | $\mu\text{g}$ | Exp. |
| $\beta^*$ | Drug loading fraction of patch | 0.0715 | 0.0686 | 0.0708 | - | Calc. |
| $k_D^*$ | Dissolution constant of polymer | 0.001 | 0.0038 | 0.0064 | cm/h | Est. |
| Th | Thickness of patch baseplate | 0.26 |  |  | cm | Calc. |
| $S$ | Total surface area of patch | 0.64 | | | $\text{cm}^2$ | Exp. |
| $S_{\text{pore}}$ | Porous surface area of skin | 0.0314 | 0.0314 | 0.0707 | $\text{cm}^2$ | Calc. |
| $V_{\text{patch}}$ | Volume of patch | 170 | | | $\mu\text{L}$ | Exp. |
| *Clinical pharmacokinetic parameters of NAL* |  |  |  |  |  |  |
| Cl | Total body clearance of NAL | 207 |  |  | L/h | [41] |
| $V_c$ | Volume of distribution of NAL in central compartment | 12.1 | | | L | [41] |
| $V_p$ | Volume of distribution of NAL in peripheral compartment | 102 | | | L | [41] |
| $Q$ | Intercompartment mass transfer rate | 284.4 | | | L/h | [41] |

Notes: 1) \*Parameters perturbed during sensitivity analyses are marked by asterisk, and values related to P3 design only were used for sensitivity analyses. 2) Assuming a square patch, patch area $S$ was estimated from pitch $w$ and needle count $N$ as: $S = \left( (\sqrt{N} + 1) \cdot w \right)^2$ . 3) $\beta$ was calculated as the ratio of drug dose to mass of patch: $\text{Dose}/(\rho \cdot V_{\text{patch}})$ , and Th was estimated as the ratio of $V_{\text{patch}}/S$ .

#### 3.6.2. Pharmacokinetic modeling

To predict the clinical pharmacokinetics of MN patch-based drug delivery, we used a two-compartment PK model [45] and simulated the plasma PK of NAL following release from the patch (**Fig. 2d)**. The following system of equations describe the PK model integrated to the drug release model via the previously calculated spline interpolant of NAL flux (*J*_NAL_(*t*)).

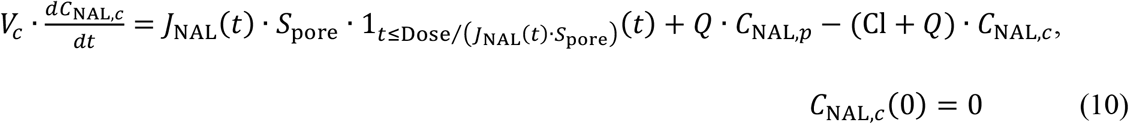

where, 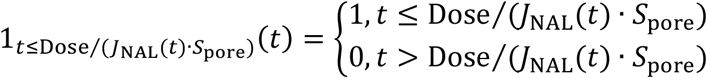

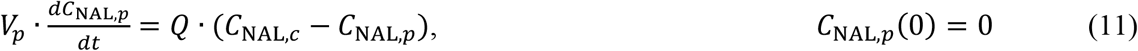

Here, *C*_NAL,*c*_ and *C*_NAL,*p*_ represent the concentration of NAL in central (i.e., plasma) and peripheral compartments, respectively; *V*_*c*_ and *V*_*p*_ are the volumes of distribution of NAL in central and peripheral compartments, respectively; *S*_pore_ represents the porous surface area of skin caused by the penetration of MNs, such that *S*_pore_ = *N* · *π* · *r*^2^; Cl is the clearance of NAL from central compartment, and *Q* is the flow rate characterizing the exchange of NAL between central and peripheral compartments. Model parameters are given in **Table 2**. The model was solved numerically as an initial value problem in MATLAB R2018a using the built-in function *ode45*.

### 3.7. Parameter analysis

To understand the effect of model parameters (physical and chemical properties of the MN patch) on plasma bioavailability of NAL, local sensitivity analysis (LSA) and global sensitivity analysis (GSA) were performed, using established methods [45–47]. For both GSA and LSA, the model parameters were perturbed within a ±50% range of the reference parameter values (given in **Table 2)**, except for parameter *w*, i.e., pitch or distance between needles (varied between -37% to +50%). Note that the lower bound for *w* was decided to ensure that pitch is never lesser than the diameter of the cone base.

In LSA, model parameters were perturbed one at a time at 1000 linearly spaced intervals between the previously defined range. For each perturbation, the PK model was solved to calculate the area under the curve (AUC_0-1h_) of the NAL plasma kinetics between 0-1 h. Note that since we are interested in rapid delivery of NAL, we ignore the long-term PK of NAL in our analysis. AUC_0-1h_ was estimated numerically via the trapezoidal method, using the built-in MATLAB function *trapz*. The qualitative nature of the relationship between NAL bioavailability and %perturbation was thus obtained.

In GSA, all model parameters were perturbed simultaneously to comprehensively evaluate and rank order the parameters for their effect on NAL bioavailability. For this, Latin hypercube sampling (LHS) [47–49] was used to obtain 1000 unique combinations of model parameters, and 10 such replicates were generated. Multivariate linear regression analysis was then performed on each replicate, and regression coefficients were determined as a measure of sensitivity index (SI) for each parameter. A distribution of regression coefficients (or SI) was obtained for each parameter from the 10 replicates, and one-way ANOVA with Tukey’s test was used to rank the parameters in terms of their sensitivity, such that a higher SI represents a greater influence on NAL bioavailability.

### 3.8. Statistical analysis

Data was analyzed using Microsoft Excel and GraphPad Prism 8.4.3.686 software package. Student’s *t* test, Shapiro Wilk test (normality evaluation), one-way ANOVA with unpaired *t* test and Welch’s correction (multiple test group comparisons) were done. Significant difference was concluded between test groups with “p” values < 0.05.

## 4. Results and discussion

Dissolvable polymeric MNs are of immense potential for the replacement of existing invasive delivery forms for NAL. As demonstrated previously by us, transdermal delivery of NAL via MNs is feasible. Dissolving polymeric needles are considered not just for the need at hand which is rapid dissolution and hence fast release but also because other MN forms such as hollow MNs are limited by features such as bore blockage with skin tissue, and the need for expensive and complicated fabrication processes such as lithography and etching [30,50]. Further, needle breakage and drug content loss are typical due to brittle fabrications. Solid needles on the other hand, though of reported usefulness for the delivery of therapeutics are associated with the need to discard sharp waste, accidental needle breakage in skin and loss of coated material [34]. Generally, with dissolvable MNs, drug is incorporated in the matrix of dissolving polymers and release is triggered on skin insertion where the polymer dissolves or disintegrates [51]. To meet the current need for NAL non-invasive and rapid delivery, the promising polymer PVP, approved by the FDA for controlled drug release alongside other uses was selected for use. It is a 20 kDa non-covalently linked dissolvable polymer with good biophysical characteristics ranging from good water solubility, tolerance to thermal treatment, great adhesion quality, biocompatibility and safety. It is cleared by the kidneys and no hazardous wastes are introduced into the environment [32,34,52]. Typical of dissolvable polymeric polymers, it also has a high drug loading capacity for hydrophilic drugs [52,53]. Numerous studies have demonstrated its effectiveness as a polymer for MN fabrication for a couple of drugs, both lipophilic and hydrophilic ranging from small molecules to biotherapeutics [34,52]. Its ability to deliver high drug flux was reported by Gao et al. in a transdermal formulation [54]. Based on the general attributes of this polymer and its successful record from numerous evaluations, our expectations following the feasibility studies on the MN based transdermal delivery of NAL using solid MNs was that for a clinical translational purpose, the development of dissolvable MNs for NAL using PVP is a veritable idea. With this approach, the limitations of solid needles are ruled out and an easy and inexpensive fabrication process which could be adaptable to large scale use of NAL MNs and as well ensure rapid delivery was envisioned. Findings on our optimized fabrication, formulation considerations and drug delivery properties are reported below.

### 4.1. Patch optimization

Various factors were observed to affect the formation of the needles ranging from the quantity of drug dissolved in polymer, polymer strength, vacuum time, and drying period. A study previously reported the effectiveness of polymer strength of about 10% for PVP. Though higher strength PVP was established to have faster dissolution rate, they were limited by skin insertion efficiency. MNs made with about 10% PVP were found to permeate the skin better [55]. The poor permeation property and weak penetration of higher strength PVP is attributed to its high-water adsorbing property [55]. Evaluations for NAL showed the efficiency of a 7.5% PVP formulation. The solubility of NAL in this PVP strength was found to be 159.08 ± 1.84 mg/ml. Considering the high drug loading capacity of this polymer and the high solubility of NAL in it, attempts were made to form high strength NAL patches with the aim of achieving fast delivery of larger amount, with large drug amount loaded in needle tips and entire patch. Based on the findings from the drug loading evaluations, 50 mg/ml formulations were eventually selected as optimal loading strength for further evaluation and optimization because for other drug concentrations evaluated (from 60 mg/ml to 150 mg/ml) a major challenge encountered was crystal formation and the formation of turbid final products with 100 mg/ml to 150 mg/ml strengths. This observation is critical as crystallization in transdermal drug formulations could affect product reliability, in that the amount expected to be delivered could be overestimated with low dose being released instead. Drug kinetics in skin becomes unpredictable as solubility could be affected and transit or drug retention or migration time is affected. Hence, poor and erratic product performance is anticipated with such high strength formulations which is undesirable from a drug formulation and clinical safety perspective. [56]. As shown in **Fig. 3 a-d**, the amount of crystals formed were observed to decrease with decreasing drug strength in dried PVP microfabrication. Their formation was thus suggested to be due to oversaturation of the 7.5% PVP solution with NAL as drying progresses with evaporation of water content. The saturation limit was thus predicted to be about 50 mg/ml NAL in 7.5% PVP in the dry state since a clear patch was achieved with this strength as seen in **Fig. 3e**. With this strength of PVP and NAL, clear needles with good integrity were obtained. Stepwise method optimization revealed that a consistent fabrication result is achievable with the steps described for the mold casting process in section 3.2 which was applied all through our studies.

**Fig. 3.**
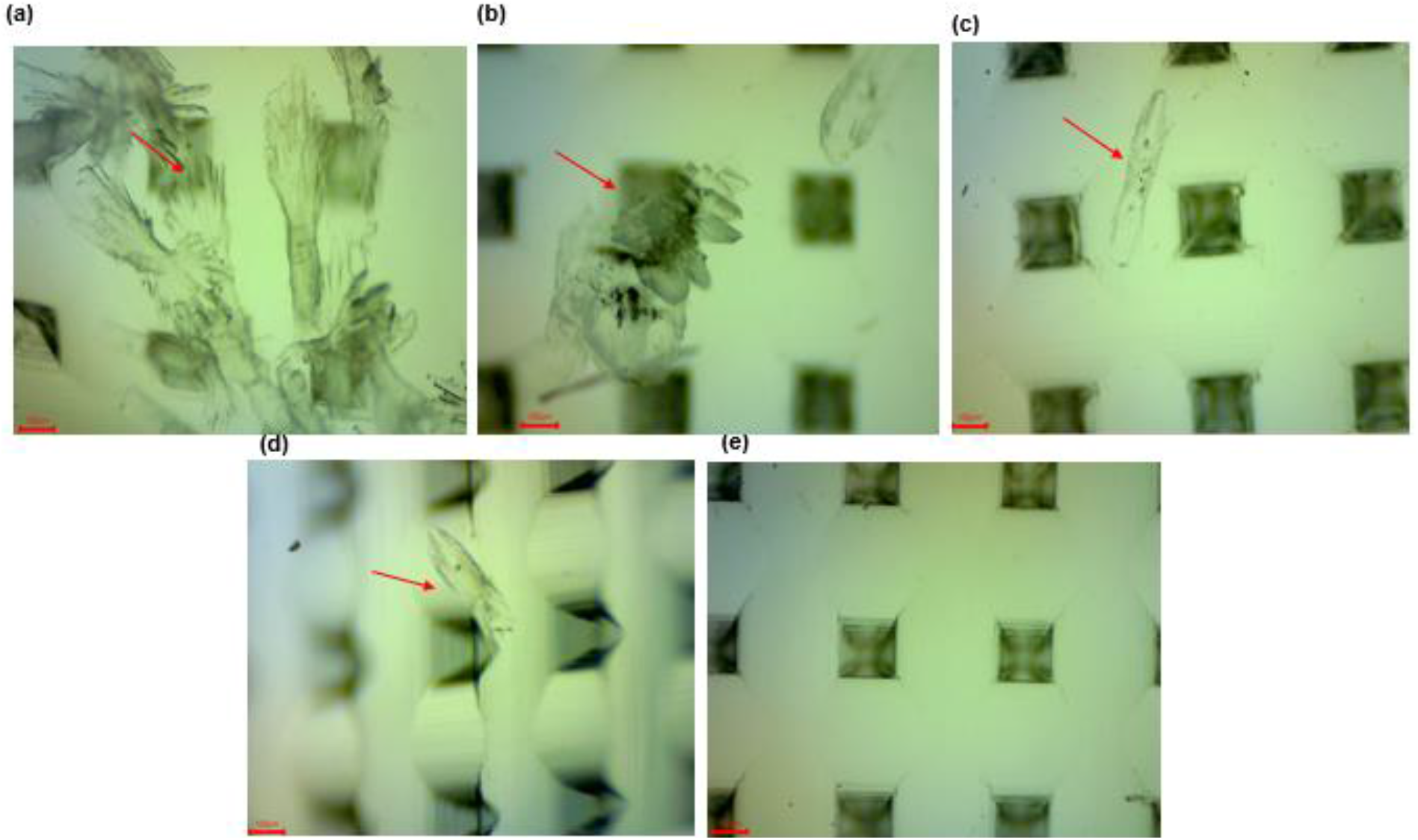
Optical microscopy of varied strength NAL microneedle patches under 40X magnification **(a)** 90 mg/ml **(b)** 80 mg/ml **(c)** 70 mg/ml **(d)** 60 mg/ml **(e)** 50 mg/ml; The red arrows depict the drug crystals

Due to vortexing/stirring to ensure uniform drug distribution and dissolution, despite sonication of drug in PVP solution, significant bubble generation is typical, and it is important to ensure complete degassing by vacuuming during fabrication as bubbles could affect needle integrity, form, and drug loading [33]. It could also influence drug release properties [31]. However, care needs to be taken regarding preventing polymer solidification and drying with prolonged handling during this degassing phase. Moreso, the final vacuuming step at a pressure of -0.6 psi for less than 2 h is also considered optimal. Excessive or prolonged pressure application was observed to cause needle base diagonal cracks which could be due to fluid being pulled down with excessive pressure. Hence, overall, attention needs to be paid to critical details and all discussed formulation considerations when being adapted to produce reliable and uncompromised products.

### 4.2. Characterization of optimized patches

#### 4.2.1. Bright Field Imaging

For the different needle dimensions, uniform needle formation was observed as shown in **Fig. 4 a-c**. MNs were conical in shape with sharp pointed tips. Morphological dimensions such as the average needle height, interspacing tip distance and base diameter for all the fabricated MNs are reported in **Table 3**. Homogeneity in the morphology across MN arrays of different dimensions was reflected by the values obtained. Moreover, the needle dimensions overall were comparable to that of master mold structures [57,58] and are within appropriate range identified in literature as what requires minimal insertion force to penetrate the skin as well as that required for painless use [58,59]. To confirm that the drug gets released at a fast rate based on the rapid dissolution property of the PVP polymer, the ability of the needle tips to dissolve within 2 min of insertion in skin was also evaluated. **Fig. 4 d-f** shows the remnants observed under the microscope following skin insertion for 2 min, which were proportionate to the height of the needles evaluated.

**Fig. 4.**
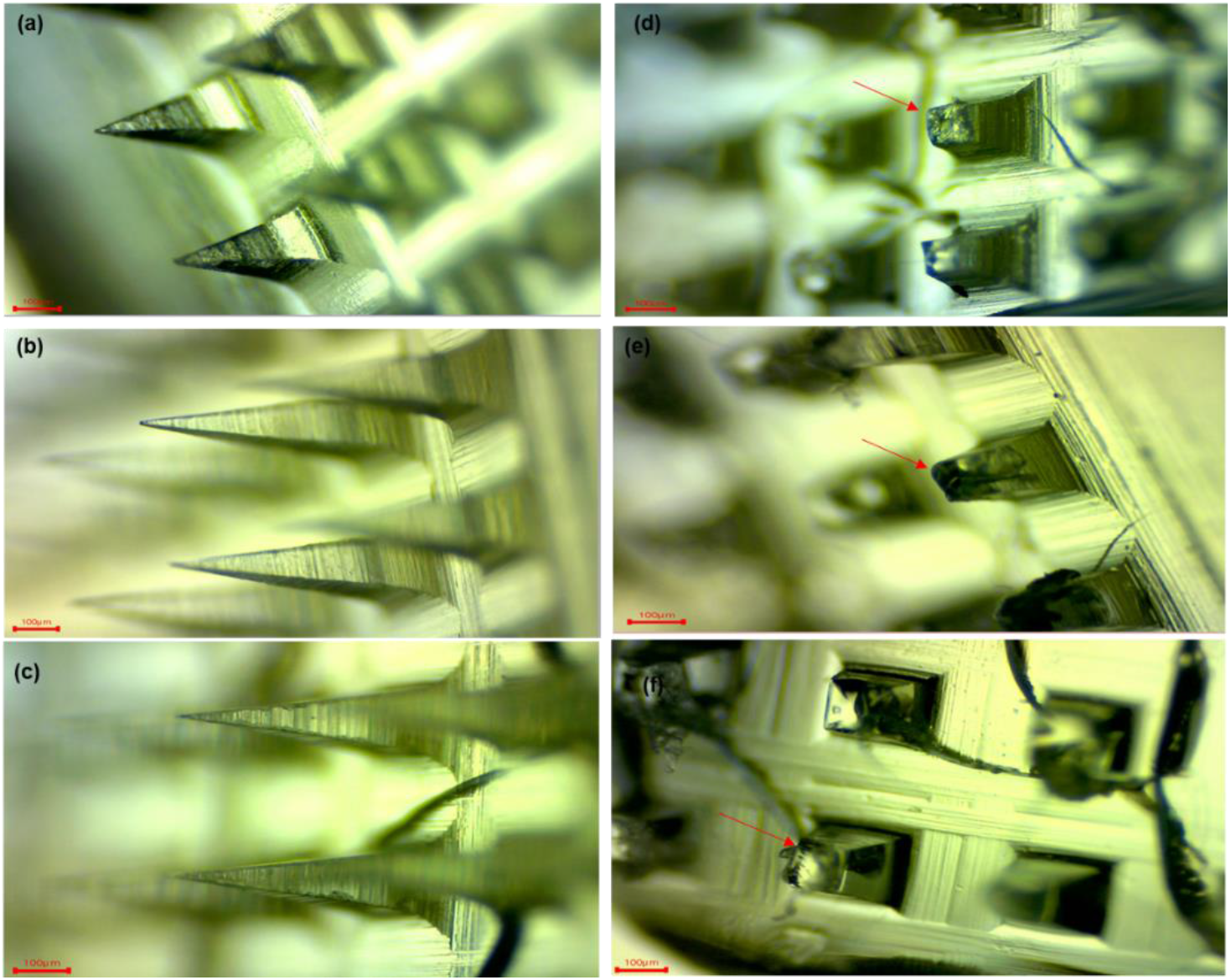
Brightfield images of the fabricated NAL patches pre-insertion for **(a)** P1 **(b)** P2 **(c)** P3 and post two min insertion in dermatomed porcine skin for **(d)** P1 **(e)** P2 **(f)** P3

**Table 3.** Dimensions of NAL-loaded PVP microneedles.

| Measurements (n = 6) | P1 | Vendors Specification | P2 | Vendors Specification | P3 | Vendors Specification |
| --- | --- | --- | --- | --- | --- | --- |
| Height ($\mu\text{m}$) | $470 \pm 1.77$ | 500 | $680 \pm 6$ | 800 | $561 \pm 2.5$ | 600 |
| Tip to tip distance (Needle Pitch ($\mu\text{m}$)) | $482 \pm 6$ | 500 | $488 \pm 3$ | 500 | $476 \pm 11$ | 500 |
| Base Diameter ($\mu\text{m}$) | $200 \pm 0.2$ | 200 | $200 \pm 0.8$ | 200 | $200 \pm 1.5$ | 200 |

#### 4.2.2. Dye Binding and skin insertion efficiency

Thumb force was used in applying needles to dermatomed porcine ear skin for 2 min. Images showing methylene blue dye retained in porated holes are shown in **Fig. 5** for all the different needle dimensions fabricated. The insertion efficiency (IE) of these was determined to be 94 ± 4.8%, 90.6 ± 1.69%, and 96 ± 1.29% for P1, P2 and P3 MNs respectively.

**Fig. 5.**
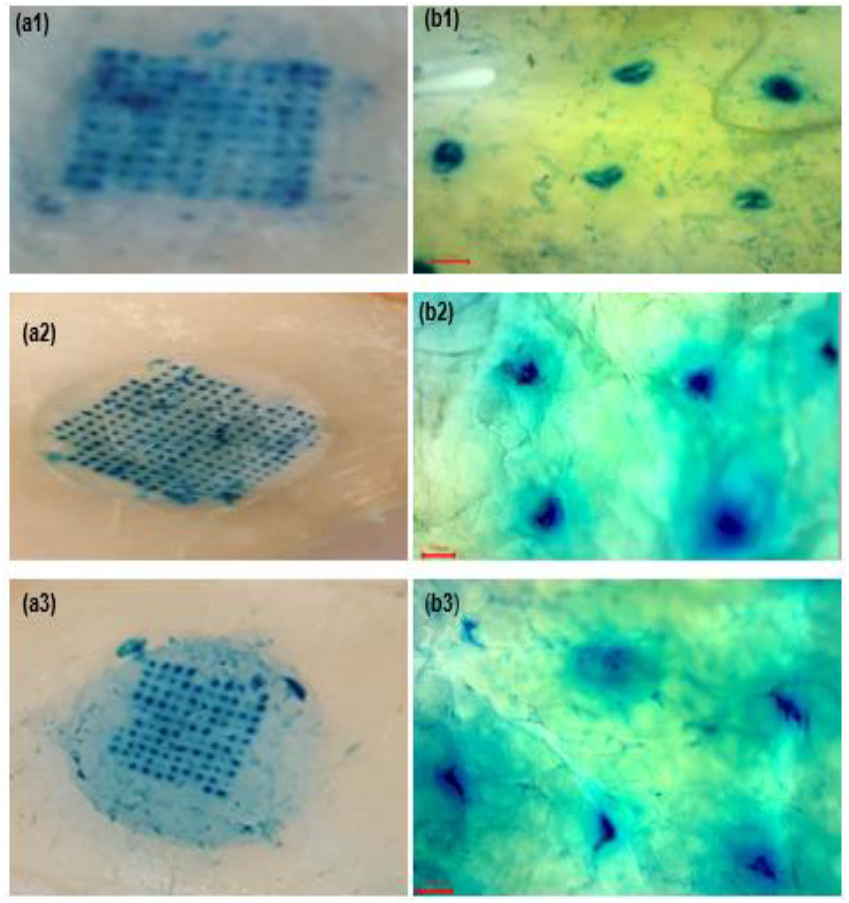
**(a)** Methylene blue dye retention on skin post needle insertion and removal (image taken with an iPhone 8 camera **(b)** Methylene blue dye retention on skin post needle insertion and removal (image taken with a brightfield microscope 40X magnification) for; (1) P1 (2) P2 (3) P3.

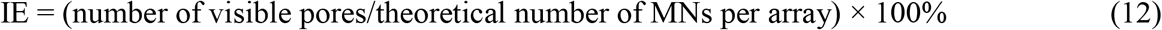

The values obtained are consistent with reports on similar dissolving microneedles made with PVP and polyvinyl alcohol which allowed a penetration efficiency of close to 100% [60,61]. The slight variations seen with insertion with different skin pieces could be related to variation in skin elasticity [62]. Another contributing factor could be the inability to maintain similar thumb force across multiple application times. In the eventuality of the clinical success of polymeric NAL MNs, the design of applicators to ensure uniform application force across skin area will be of good use [63]. The retention of dye in the pores indicates efficient skin penetration and the formation of microchannels for the passage or diffusion of drugs. This also further indicates that the needles have sufficient mechanical strength to withstand the insertion force applied manually [21].

#### 4.2.3. Confocal laser scanning microscopy

Confocal microscopy study further confirmed the capability of formed needles to porate skin and to allow dye to rapidly diffuse through created microchannels. Within 1 min of applying fluorescein as a fluorescent dye for imaging, a total dye diffusion length of 150 ± 4.08 µm was observed for the 800 µm NAL MN, 130 ± 2.5 µm and 80 ± 0.1 µm for the 600 µm and 500 µm MNs, respectively as shown in **Figs. 6a-c**. There is evidence to show that they all produced microchannels sufficient for the transport of drug. As noted by Marmato et al., MNs usually penetrate 10-30% of their actual length in skin. This thus explains the different diffusion depth observed with confocal microscopy for the different needles which falls within this range. Typically, it is expected that an indent is initially created by needles in skin which dependent on pressure applied gets to break into the skin after a while during application [58,64]. The depth of channel created then varies with the amount of pressure applied, skin viscoelasticity and integrity [58]. Also, needle height is seen to correlate with diffusion path with 800 µm needles showing deeper dye diffusion depth compared to the rest within 1 min. The measured dye diffusion depth for each of the dimensions of MNs are consistent with observations from previous studies involving dissolving MNs. Liu et al. reported a diffusion length of 200-400 µm following application of 500 µm PVP MNs preloaded with fluorescein isothiocyanate for 15 and 30 min [65]. Similarly, histological evaluation of the actual depth created by hyaluronic acid microneedles via histological sectioning of 500 µm MN porated skin recorded a depth of about 47.3 to 99.3 µm [66]. Also, for 650 µm PVP MNs, a study reported an insertion depth of 200 µm [30]. Therefore, the conclusion reached is that the PVP based polymeric MNs for NAL are of good performance considering the evidence of channel creation in skin.

**Fig. 6.**
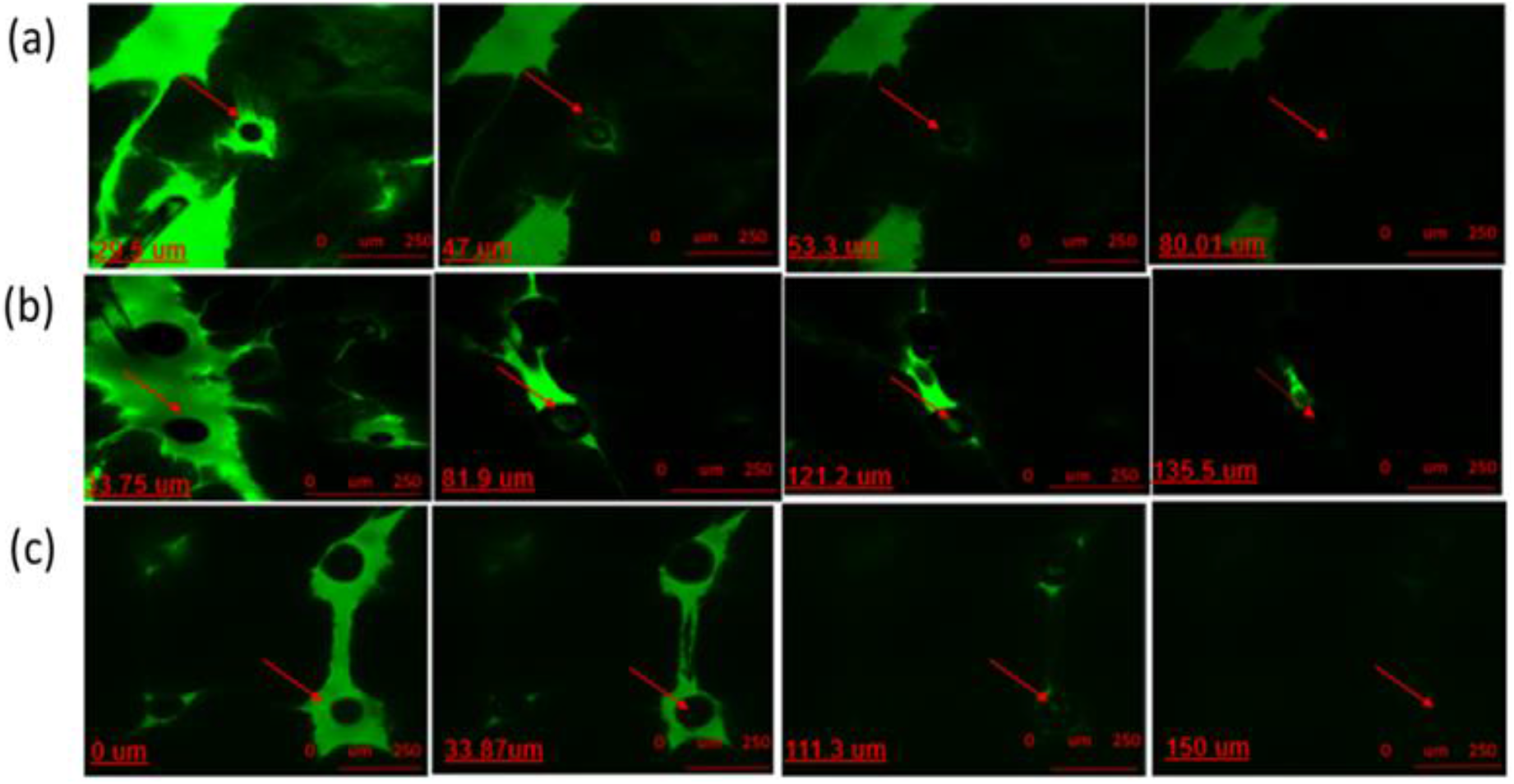
Confocal microscopy images depicting the depth of microchannels created by treatment with **(a)** P1 **(b)** P2 and **(c)** P3.

#### 4.2.4. Drug content

For all drug formulations, it is essential to analyze the quantity of drug in final formulation to have an estimate of the exact amount being dosed. Various factors ranging from spillage during the mold filling step, drug instability in formulation to crystallization could significantly impact final drug content. For all microfabricated needles, the theoretical NAL content per mold is 22.5 mg. The experimentally determined percentage drug content was 83.2 ± 1.06% (17.89 ± 0.23 mg), 80.02 ± 3.6% (17.2 ± 0.77 mg), and 82.8 ± 2.3% (17.8 ± 1.01 mg) for P1, P2 and P3 respectively. In this study, explanations for the almost 20% drug loss in the optimized formulations is attributed to the spill of drug in polymer solution from micro-molds during the mold casting process, particularly, the degassing and vacuuming phase. Fabrication under controlled and robust conditions are anticipated to minimize such loss. However, the reported drug contents are considered to still be within narrow limits from the theoretical amount of drug envisaged to be incorporated in MN arrays. Also, uniformity in drug content across different arrays are seen with the insignificant variation seen across multiple arrays SE (n=4).

### 4.3. *In vitro* permeation study

#### 4.3.1. Passive permeation of NAL hydrochloride

Few studies have evaluated the transdermal delivery of NAL. For what it is worth this route is established by these studies as a preferred route for the delivery of NAL considering the numerous advantages users of this drug could benefit from. Passive permeation of NAL via skin was shown by these research studies to be quite deficient and this limitation is established to be due to the hydrophilic property of NAL which limits its capability to diffuse through the lipid bilayer of skin [13,67]. Similar to these previous findings, we also observed no significant drug flux with passive delivery (1.53 ± 0.83 µg/sq.cm/h over 24 h) as shown in **Fig. 7a**. This data compares to that on passive delivery from our previous study for which a flux of 1.75 ± 0.25 µg/sq.cm/h was recorded [13]. In this current study, no significant drug flux was seen in the first 1 h of passive delivery and a lag time of 62.057 ± 4.05 min was also observed. Hence, despite the presented advantages, such as minimal invasiveness, sustained delivery, improved bioavailability and the provision of easy options to firsthand care givers for the management of NAL emergencies, the utmost desire remains rapid release, to avoid loss of patients prior to the onset of some of the benefits associated to the route. This requirement is justified by the need for urgent recovery when patients present with respiratory depression and the need for very rapid stabilization with NAL [68]. No form of delivery indeed beats the bolus delivery possible with the IV route, however the development of delivery systems with properties quite similar to IV delivery or at least close enough to that seen with IM, SC or IN dosing are considerable options.

**Fig. 7.**
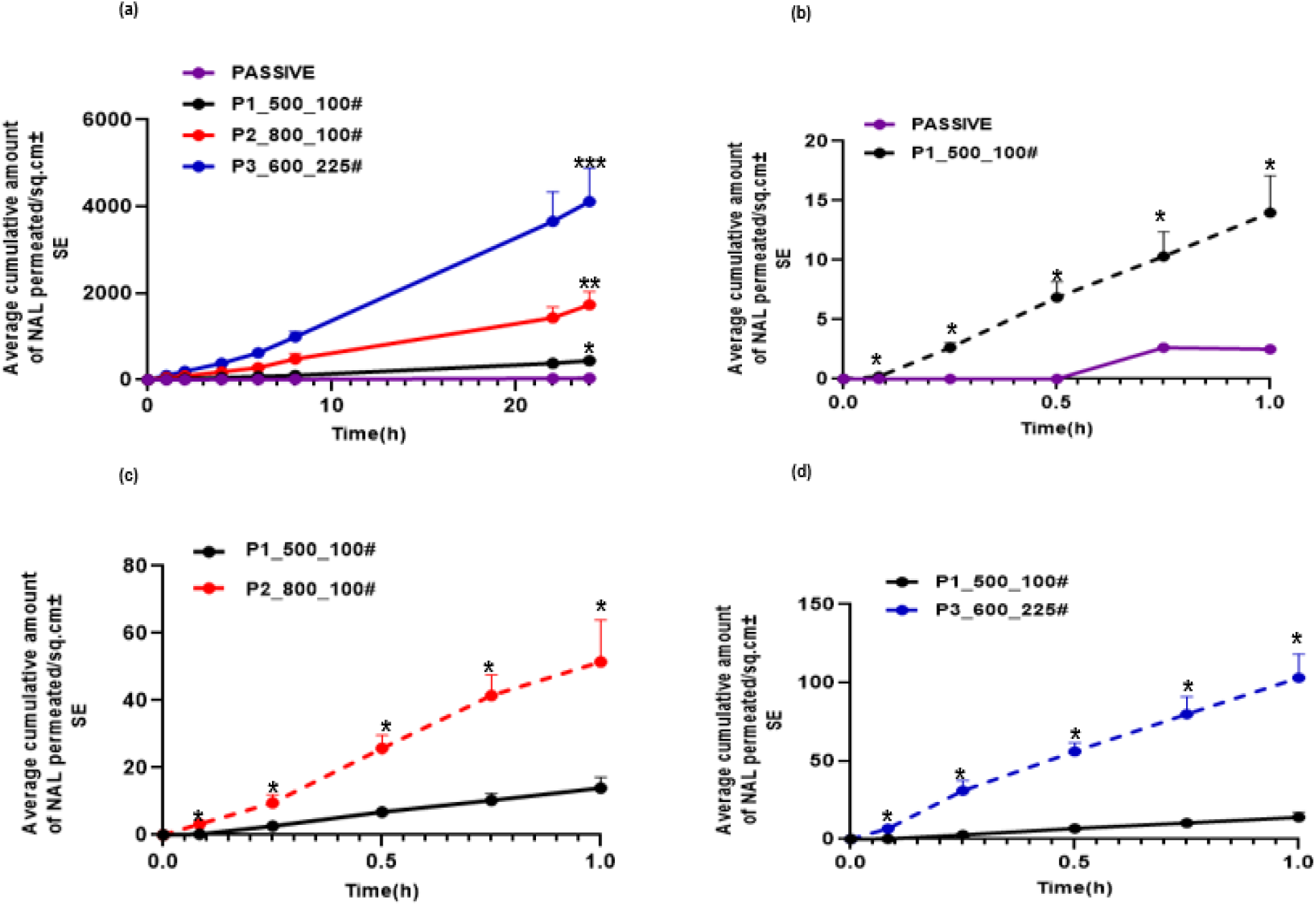
**(a)** 24 h permeation profile of passive delivery and all fabricated MNs **(b)** Average cumulative amount permeated per sq.cm over 1 h of NAL, comparing passive delivery and P1 **(c)** P1 and P2 **(d)** P1 and P3 (p<0.05, *= significant difference at individual time points)

#### 4.3.2. Effect of MNs: Passive Vs P1

As a significant improvement over the passive permeation data from the current study (p<0.05), as shown in **Fig. 7a**, the fabricated PVP based MN, P1 produced an average drug flux of 18.71 ± 3.24 µg/sq.cm/h. and an average cumulative amount of 440.14 ± 29.93 µg/sq.cm over 24 h. This improvement is expected with evidence from the dye binding study and confocal imaging of dye diffusion depth of MN array confirming the creation of transport channels for drug to pass through. Further, evidence of sustained delivery is seen and the need for multiple injections to stabilize patients is expected to be eliminated. As shown in all previous studies, the major obstacle with the transdermal delivery of NAL has been the lag time to drug delivery which ranges between 1.6-11.5 h [13,20,67]. For a transdermal system being considered for use in opioid emergencies, rapid delivery in the first one hour of drug application is considered very critical to enable quick reversal of opioid complications. As shown in **Fig. 7b**, the treatment of dermatomed porcine ear skin resulted in higher permeation (p<0.05) of NAL with P1 in the first 1 h at the respective time points over zero permeation amount seen with passive permeation at these time points. The average flux over the first 1 h with P1 was also 15.09 ±7.68 µg/sq.cm/h. As was envisaged prior this study, the overall data with P1 reflects significant permeation enhancement and a reduction in lag time to 5 min (∼ 90%) (p<0.05) over what was observed with the passive delivery of NAL. Also, compared to the 8 min delivery lag time previously reported in preliminary studies from our lab with the use of solid MNs [13], the 3 min difference seen in this current study with dissolvable MNs is considered significant and should make some difference with the onset of drug effect when used *in vivo*. Further, since the drug is loaded in the needle itself, time taken to initially apply solid needles before applying drug is ruled out as drug dissolves in skin with insertion and there is no hazardous waste generated with this. Moreso, the risk of needle breakage in skin is eliminated. The applicability of these MNs for the rapid delivery of NAL in clinical overdose presentations is thus evident.

#### 4.3.3. Effect of MN length: P1 Vs P2

Interest in options to improve on drug flux particularly in the first one hour led to the evaluation of the effect of increasing MN length on the permeation of NAL. The patch P2 (Table 1) consisted of an array of MNs with length of 800 µm (300 µm greater than P1), while all other fabrication parameters held constant. As shown in **Fig. 7a**, the overall permeation profile of P2 over 24 h was significantly higher (p<0.05) than that of passive delivery and P1. Also, as shown in **Fig. 7c**, an exciting observation for P2, in the first 1 h was a significant increase (p<0.05) in drug flux and amount permeated across all individual time points evaluated compared to what was obtainable with P1. The average drug flux and amount permeated in 1 h with P2 were 60.44 ± 22.6568 µg/sq.cm/h and 51.5 ± 12.41 µg/sq.cm respectively (p<0.05), which are evidently higher than the values recorded with P1. Data from the confocal study with the creation of deeper diffusion depth by P2 explains the difference seen reflecting the creation of deeper channels by longer needles and hence higher permeation in same time points. Data from this study compares to that from similar studies where needle length has been shown to correlate with the depth of permeation and amount permeated [69]. In the previous study by us, 250 µm MNs produced shallow channels in comparison to 500 µm MNs and this as well reflected a reduced permeation in comparison with the former, with longer length [13]. Banga et al. also made a similar observation with maltose MNs, for which depth of channels created was found to increase with increasing MN length and this as well reflected in the amount of loaded drug permeated [58].

#### 4.3.4. Effect of MN length and density: P1 Vs P3

The effect of increasing needle density i.e., number of MNs per unit area with a slight increase in MN length on drug flux and amount permeated was also evaluated. As shown in **Fig. 7d**, P3 enabled much more significant amount of permeation compared to P1 over 24 h (p<0.05). It was observed as well that within the first 1 h of application on dermatomed porcine skin the average flux for P3 within 1 h was 102.83 ± 32.34 µg/sq.cm/h which is almost double that seen with P2 and about 7-fold the flux seen with P1 (p<0.05, **Fig. 8)**. Permeation increase with increased needle density is consistent with reports from previous studies where increase in permeation with increase in MN density is attributed to generation of more holes in skin [70,71]. Though some variations were seen across samples as is evident in the SE of measurements, Lahiji et al. ascribes variations seen across sampling with skin pieces involving MNs to the difference in skin elasticity across skin pieces used in the studies as well as the amount of hair retained on skin post shaving which could influence penetration depth for each MN on an array across samples [62]. Also, as previously discussed variation in pressure at the point of needle application could result in varied results across skin samples [58].

**Fig. 8.**
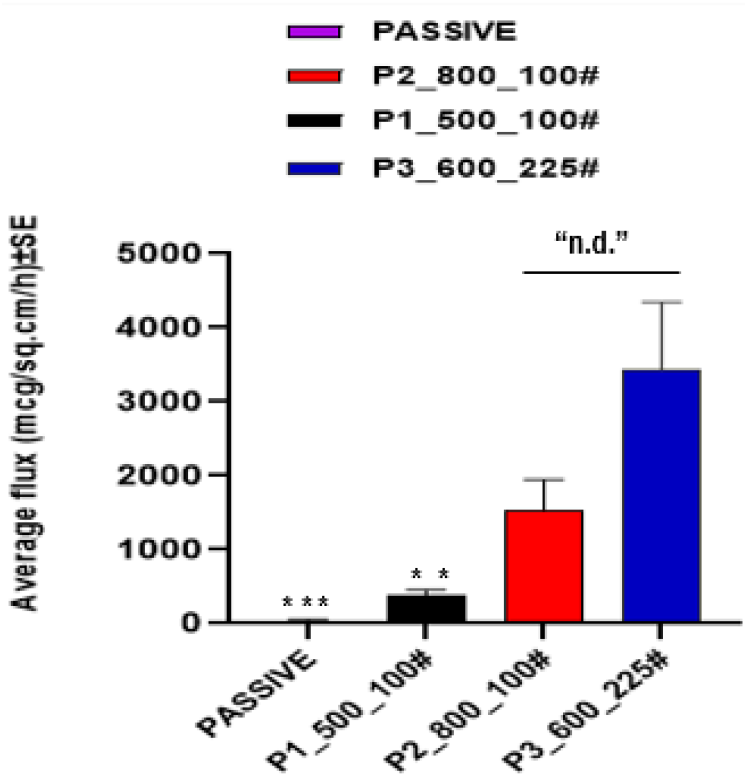
*In vitro* over 1 h permeation flux of NAL with passive delivery, P1, P2, and P3 (p<0.05 ***=significantly higher than passive, **=significantly higher than passive and P1, “n.d.” no significant difference

The results from this study evaluating the influence of MN length and density on NAL permeation/flux through skin indicates that both factors significantly impact the amount of drug deliverable. Increasing MN density shows more promising influence with the same strength of drug (50 mg/ml). Therefore, it can be concluded that creating more channels on skin for drug to diffuse through a larger area ensures a higher drug diffusion rate compared to increasing channel depth in skin using longer length needles. This observation was as well seen with the data observed with confocal imaging studies, though longer length needles produced deeper diffusion length within 1 min of dye application, the difference seen with the slightly shorter length larger array needle wasn’t so significant. Again, this could be attributable to skin elasticity, preventing the full length of the longer needle from getting into skin. As seen with the 2 min post dye insertion images for P2, longer remnants post patch removal is seen **Fig. 4e**. This indicates that there might be a limit to the yielding capacity of skin elasticity as MN goes in perpendicularly. Overall, both the *in vitro* permeation studies and needle characterization studies confirm the effectiveness of the polymeric needles for porating the skin to create transient channels for the diffusion of drug compared to an intact skin surface where the hydrophilic drug is limited from crossing the rate limiting barrier SC. The feasibility of manipulating patch array density and length to achieve significant therapeutic doses of the drug is also seen.

### 4.4. Mathematical modeling and simulation

#### 4.4.1. Model development and clinical pharmacokinetic predictions

The drug release model (Eqs. 8, 9) was fit to *in vitro* drug release kinetics data for the three patch designs studied here (**Fig. 9a)**, and the unknown model parameter *k*_*D*_ was thus estimated for each design (**Table 2)**. Since the region of interest for application in opioid overdose treatment is the first few minutes to an hour, the model was thus calibrated to reproduce the short-term (up to 1 h) experimental behavior only. As shown in **Fig. 9a**, the numerical solution of the model is in good agreement with the data for all three patch designs, as also indicated by the Pearson correlation coefficient *R>*0.99.

**Fig 9.**
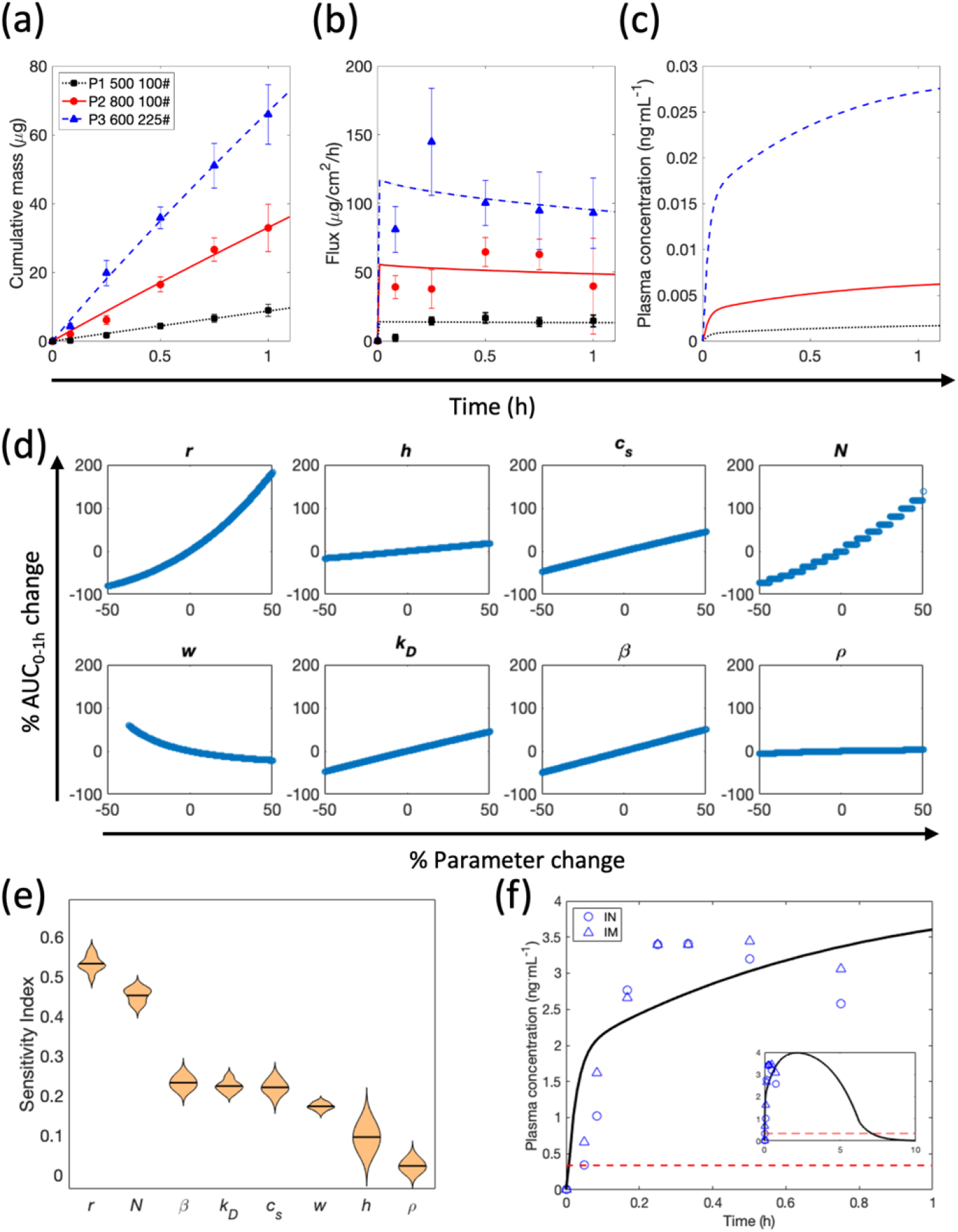
Modeling and simulation analysis. **(a)** Nonlinear regression analysis of Eqs. 8, 9 to *in vitro* data for the three patch designs investigated in this study. (**b)** Spline interpolation to the predicted flux kinetics of NAL compared to experimentally calculated flux for the three patches. Data represents mean±SEM. (**c)** Clinical predictions of NAL plasma pharmacokinetics from the two-compartment model for the three patch designs. (**d)** Local sensitivity analysis showing qualitative relationships between %AUC_0-1h_ change and % perturbation in parameter values for different model parameters. (**e)** Global sensitivity analysis-derived rank ordering of model parameters in terms of their sensitivity indices (SI) reflecting their effect on plasma AUC_0-1h_ of NAL. Greater the SI, higher the parameter sensitivity. (**f)** Optimization of the P3 patch design to match the plasma pharmacokinetics of NAL obtained through commercial intranasal (IN) and intramuscular (IM) devices. Inset shows the long-term behavior of the optimized patch and dashed red line represents the minimum effective concentration of NAL.

From the cumulative mass kinetics profile, NAL flux (*J*_NAL_(*t*)) was calculated and spline function was fit to it, which showed good qualitative agreement with the experimentally calculated flux (**Fig. 9b)**. The spline interpolant *J*_NAL_(*t*) was then used to predict the plasma PK of NAL for all three designs using a two-compartment PK model (Eqs. 10, 11), and as shown in **Fig. 9c**, the P3 design which produced the highest flux, led to the greatest bioavailability of NAL, followed by P2 and P1 patches. However, the plasma concentration achieved with the current designs is much lower than the pharmacologically effective NAL concentration value of ∼0.34 ng/mL [13], and thus optimization of patch design is necessary to achieve the minimal effective concentration, or to match the performance of FDA-approved devices for NAL delivery.

#### 4.4.2. Model-guided patch optimization

To understand the impact of model parameters on drug delivery and plasma bioavailability of NAL, we performed LSA and GSA with eight model parameters and assessed the effect of parameter perturbations on area under the plasma concentration kinetics curves during the first 1 h (AUC_0-1h_). As shown in **Fig. 9d**, parameter perturbations between ±50% of the baseline values led to the greatest change in AUC_0-1h_ in case of the MN cone radius *r* and MN count *N*, with both parameters showing a positive correlation with AUC_0-1h._ The remaining parameters showed only mildly positive correlations with AUC_0-1h_, except for patch material density *ρ* that appears to have no impact on NAL bioavailability and pitch *w* that was mildly negatively correlated to AUC_0-1h_. Of note, MN height *h* only mildly affects AUC_0-1h_, which can be attributed to the model assumption that needles penetrate deep enough to be close to the microvasculature in the dermis of the skin. However, future analysis is necessary to incorporate the effects of mechanical strength of MNs and skin elasticity to estimate the true depth of MN penetration for a given height *h*. The polymer-related parameters, i.e., solubility *c*_*s*_ and dissolution constant *k*_*D*_ also positively affect AUC_0-1h_, thereby providing opportunities to enhance NAL bioavailability by modifying polymer composition. Also, drug loading fraction *β* is mildly positively correlated with AUC_0-1h_, indicating that higher drug loading fraction can be considered to enhance NAL bioavailability, as long as it does not negatively impact MN strength. These observations suggest that increasing MN count and MN base diameter can strongly, positively impact NAL bioavailability, which can further be improved by keeping a shorter pitch, i.e. a higher MN density, as also corroborated by our experimental findings.

We further confirmed these observations through GSA, where model parameters were simultaneously perturbed to evaluate their impact on NAL bioavailability and also to understand the impact of potential parameter interactions. As shown in **Fig. 9e**, parameter ranking order from GSA validates the findings of LSA indicating that *r* and *N* are the most potent parameters in affecting NAL AUC_0-1h_.

As a result, we used these two parameters to optimize our best performing patch design (P3) to match the clinical PK of NAL obtained via FDA-approved intranasal (IN) and intramuscular (IM) devices [72]. As shown in **Fig. 9f**, the PK model was fit to the combined IN and IM clinical data to optimize parameters *r* and *N*, while all other model parameters were fixed at the baseline values for the P3 patch. Our results indicate that the best fit of model predictions with clinical data was obtained for *r* = 0.0238 cm and *N* = 2704 needles (i.e., 52×52 array). Thus, increasing the P3 patch to an area *S* of ∼6.4 cm^2^ with MN base dimensions to achieve *S*_pore_ ∼4.8 cm^2^, while keeping the other parameters unchanged can reproduce the clinical PK profile of NAL achieved through industry standard devices. Note that *β* was held constant at 0.0708 (i.e. ∼7 % drug loading by weight) in our optimization, thus the loaded dose in the optimized patch is ∼185 mg.

Of note, as shown in the inset in **Fig. 9f**, not only does the optimized patch design match the initial clinical performance of gold-standard devices, it also produces a long-lasting effect due to plasma concentration levels of NAL remaining above the effective concentration for up to ∼7 h post administration. It can thus be inferred that increasing MN count while also increasing MN base diameter is an effective way to enhance the porous fraction of the skin under impact by the patch, thereby leading to enhancement in drug flux and plasma bioavailability.

## 5. Conclusion

NAL dissolvable polymeric PVP based MNs were effective in creating microchannels in skin to increase drug diffusion. This was evident by higher flux rate achieved compared to passive permeation. The mechanical strength, needle insertion efficiency and uniformity of content for different batches of needles earmark the reliability of the microfabrication process applied and the success of applying PVP based NAL MNs in clinical settings is predicted. A major finding from this study was the reduction in lag time seen with NAL PVP based MNs to about 5 min compared to the lag time of previously evaluated transdermal systems for NAL which would make a major difference on the onset of effect when applied in emergency opioid situations. The association between increase in needle length and needle density (No. of needle/per unit area) on drug permeation was established. Of note, through mathematical modeling and simulations we optimized the patch design to reproduce clinical PK of NAL obtained through FDA-approved intranasal and intramuscular devices, which helped us identify the significance of needle base diameter and needle count in governing the bioavailability of NAL. Ultimately, the applicability of rapidly dissolving NAL PVP based MN for either the stabilization of opioid overdose patients or for maintenance therapy following stabilization to eliminate opioid induced side effects such as pruritus and constipation are foreseen.

## Data Availability

All data produced in the present study are available upon reasonable request to the authors

## Abbreviations

CLSM: Confocal laser scanning microscopy
FDA: Food and Drug Administration
GSA: Global sensitivity analysis
HIV/AIDS: Human immune deficiency syndrome/Acquired immune deficiency syndrome
HPLC: High performance liquid chromatography
NAL: Naloxone
NIDA: National Institute on Drug Abuse
MN: Microneedle
IE: Insertion efficiency
IN: Intranasal
IM: Intramuscular
IV: Intravenous
LSA: Local sensitivity analysis
GSA: Global sensitivity analysis
PBS: Phosphate buffered saline
PDMS: Polydimethylsiloxane
PK: Pharmacokinetics
PVP: Polyvinylpyrrolidone
SC: Stratum corneum
TFA: Trifluoroacetic acid

## 6. Acknowledgements

Prashant Dogra and Maria J. Peláez thank Dr. Javier Ruiz Ramírez for helpful scientific discussions with mathematical model development. Ashana Puri would like to thank Dr. Hiep Nguyen for useful discussions pertaining to microneedle fabrication protocol.

## Statements and declarations

### Author contributions

Conceptualization and design of project was performed by: [Ashana Puri], [Akeemat O. Tijani]; Methodology was designed by: [Ashana Puri], [Akeemat O. Tijani], [Prashant Dogra], [Maria J. Peláez]; Formal analysis and investigation was performed by: [Ashana Puri], [Tijani Akeemat], [Prashant Dogra], [Maria J. Peláez]; Writing - original draft preparation was performed by: [Akeemat O. Tijani], [Ashana Puri], [Prashant Dogra]; Writing - review and editing was performed by: [Akeemat O. Tijani], [Ashana Puri], [Prashant Dogra], [Maria J. Peláez], [Zhihui Wang], [Vittorio Cristini]; Funding acquisition was by: [Ashana Puri], [Prashant Dogra], [Zhihui Wang], [Vittorio Cristini]; Resources were made available by: [Ashana Puri]; Supervision was done by: [Ashana Puri]. All authors read and approved the final manuscript.

### Data availability

The datasets generated during and/or analyzed during the current study are available from the corresponding author on reasonable request.

